# Healthcare utilization and clinical characteristics of genetic epilepsy syndromes: a longitudinal case-control study of electronic health records

**DOI:** 10.1101/2023.05.27.23290634

**Authors:** Christian M Boßelmann, Alina Ivaniuk, Mark St John, Sara C Taylor, Gokul Krishnaswamy, Alex Milinovich, Costin Leu, Ajay Gupta, Elia M Pestana-Knight, Imad Najm, Dennis Lal

## Abstract

**Background:** Understanding disease progression, age-specific comorbidities, medical treatment patterns, and unmet needs can help improve the care pathway of individuals with rare genetic epilepsies. A matched longitudinal cohort study has not been performed for these variables from childhood to adolescence across the whole phenome.

**Methods:** We identified individuals with likely genetic and non-genetic epilepsy syndromes and onset at ages 0-5 years by linkage across the Cleveland Clinic Health System. We used natural language processing to extract medical terms and procedures from longitudinal electronic health records (EHR) and tested for cross-sectional and temporal associations with genetic epilepsies.

**Findings:** We identified 503 individuals with genetic epilepsy syndromes and matched controls with epilepsy that did not receive genetic testing. The median age at the first encounter was 0·1 years, 7·9 years at the last encounter, and the mean duration of follow-up was 8·2 years. We extracted 188,295 Unified Medical Language System (UMLS) annotations for statistical analysis across 9,659 encounters. Individuals with genetic epilepsy syndromes received an earlier epilepsy diagnosis and had more frequent and complex encounters with the healthcare system. Notably, the highest enrichment of encounters compared to the non-genetic groups was found during the transition from paediatric to adult care. Our computational approach could validate established comorbidities of genetic epilepsies, such as behavioural abnormality and intellectual disability. We also revealed novel associations for genitourinary abnormalities (OR 1·91, 95% CI: 1·66-2·19, p = 2·39×10^-19^) linked to a spectrum of underrecognized genetic syndromes.

**Interpretation:** This study identified novel features associated with the likelihood of a genetic epilepsy syndrome and quantified the healthcare utilization of genetic epilepsies compared to matched controls with epilepsy who did not receive genetic testing. Our results strongly recommend early genetic testing to stratify individuals into specialized care paths, thus improving the clinical management of people with genetic epilepsies.

**Funding:** Not applicable.

## Introduction

Many forms of epilepsy are likely to have a genetic aetiology, ranging from rare *de novo* monogenic syndromes like developmental and epileptic encephalopathies (DEE) to polygenic burden in common focal and generalized epilepsies.^1,2^ Overall, >140 epilepsy-associated genes have been identified.^3^ While individually rare, the annual incidence of genetic epilepsies is estimated to be 1 per 2120 live births.^3^ These syndromes were historically defined by careful observation of the key clinical features of small cohorts. More recently, electronic health records (EHR) have been applied to scale this discovery process to the large amount of data available today. Standardized vocabularies and ontological reasoning have enabled and partially addressed the inherent limitations of using large-scale real-world data.^5,6^ Deep quantitative phenotypic analysis has greatly enhanced our understanding of the clinical spectrum of disorders related to variants in *SCN2A*^7^, *STXBP1*^8^, and others. Longitudinal approaches have examined the disease trajectories of rare syndromes to identify age-dependent patterns in their clinical features across thousands of patient years.^9,10^

While previous work has focused on deep data analysis from individuals with variants in known epilepsy-related genes, the practical implications for a larger and more general population sample remain unclear. Individuals with childhood-onset genetic epilepsy syndromes are known to have heterogeneous clinical features^11^ and are affected by high rates of psychiatric and somatic comorbidities.^12^ Their disease progression from childhood to adolescence and the impact on healthcare resource utilization and medical treatment are poorly understood.

Genetic testing is vital to address their unmet medical needs, as it facilitates a timely diagnosis, informs clinical management, and enables candidate precision therapies or clinical-trial readiness.^13^ Certain clinical features increase the pre-test probability of positive genetic testing.^14^ Hence, the Genetics Commission of the International League Against Epilepsy (ILAE) recommends genetic testing in cases with additional symptoms, including intellectual disability, autism, dysmorphology, and others.^15^ Identifying clinical features that are independently associated with genetic epilepsy syndromes may therefore improve patient selection for testing.

Here, we conducted a case-control cross-sectional and longitudinal study on EHR data from individuals with known or likely genetic epilepsy syndromes against matched controls with epilepsy across a large healthcare network. We set out to describe the disease progression, comorbidities, and medical treatment of individuals with likely genetic epilepsy syndromes. Our data-driven whole-phenome approach identifies novel clinical features predictive of genetic epilepsy syndromes and highlights the unmet medical needs of these individuals.

## Methods

### Setting and Participants

This study was carried out at the main campus and 14 north-eastern Ohio affiliate hospitals of the Cleveland Clinic Health System of the Cleveland Clinic Foundation. Electronic health records were queried for entries between 01/01/1998 and 31/01/2023. The study site is a Level 4 Adult and Paediatric Epilepsy Centre accredited by the National Association of Epilepsy Centers (NAEC). We chose the setting of a large healthcare system network to reduce the impact of single providers, enable data sharing across sites, and benefit from standardized professional guidelines and coding practices. All sites used Epic electronic medical records (Epic Systems Corporation, WI, USA).

Eligibility criteria to identify epilepsy cases were: i) Any International Classification of Diseases, Tenth Revision, Clinical Modification (ICD-10-CM) code G40-(“Epilepsy and recurrent seizures”) or ICD-9 code 345.*; ii) Any Current Procedural Terminology (CPT) code for electroencephalography (EEG); iii) Age 0-5 years at the time of diagnosis (first billing code for epilepsy). Eligibility criteria were based on a systematic meta-review on the accuracy of using administrative healthcare data to identify epilepsy cases, where the positive predictive value and sensitivity of nine validation studies in the US ranged from 32·7 – 96·0% and 12·2 – 97·3%, respectively.^16^ We chose strict cohort definitions based on two rationales: i) Participants who had received CPT codes for EEG may be more likely to have been diagnosed within the healthcare system, increasing length and depth of follow-up; ii) Participants should be strongly enriched for epilepsy while removing those with unclear diagnoses such as convulsions or syncope (i.e., high precision at the cost of sensitivity).

Participants were then stratified into case-control groups for further analysis. Likely genetic individuals had ≥ 1 order for any genetic testing and a match for a custom natural language processing (NLP) algorithm (Table S1). Likely non-genetic individuals had neither. Additional individuals that fulfilled the eligibilitiy criteria were identified by ICD-10 codes for monogenic syndromes, including tuberous sclerosis complex (ICD-10 85·1, n = 17), Cyclin-Dependent Kinase-Like 5 Deficiency Disorder (CDKL5-DD, ICD-10 G40·42, n = 11), or Dravet syndrome (ICD-10 G40·834, n = 13). For additional validation, we implemented PheIndex, a recently developed algorithm to identify individuals with rare genetic disorders, and found a strong correlation with our group labels (Figure S1).^17^

For the control group, we applied three matching criteria with the following rationales: i) Sex, as several genetic epilepsy syndromes and their comorbidities have sex-dependent phenotypic features; ii) Median age, to control for differences in age-dependent longitudinal phenotypes and changes in billing or coding practices; iii) Self-reported ancestry, to minimize systematic bias of our genetic risk estimates by ancestry-dependent population substructure. Matching was done by propensity score matching with scores estimated by a generalized linear model followed by nearest-neighbour matching at the default 1:1 ratio.^18^ After matching, the final study cohort consisted of 503 individuals. Due to the nature of the retrospective EHR-based study design, information on individuals lost to follow-up was unavailable.

### Variables

The investigators had full access to the database population used to create the study population. Dataset construction, cleaning, and person-level linkage across the three databases (Figure 1) were carried out as previously described.^5^ The Research Data Warehouse at the Cleveland Clinic is an in-house relational database that maps Unified Medical Language System (UMLS, release 2022AA) concepts to integrate and standardize clinical data. This process includes automatic source code matching (2011 ICD-9-CM, 2023 ICD-10-CM, CPT 2021), exact or fuzzy text matching to raw clinical notes (Apache cTAKES), and manual mapping. More than 70% of data is mapped automatically, and the system has been previously validated across a wide range of use cases^5^. This procedure resulted in a list of UMLS concept annotations for each person with epilepsy at every encounter. Duplicates were removed, and concepts were grouped if their encounters occurred within one month of each other (as codes generated during billing, lab results, or late documentation were assigned different dates). These concepts were then mapped to Human Phenotype Ontology (HPO, v2023-01-27) terms, a standardized vocabulary of phenotypic features.^6^ The use of the HPO as a phenotyping algorithm has been previously validated, and the process of propagating sets of terms to enable ontological reasoning has been previously described (Figure S2).^7,8^ The comprehensive ontological system of the UMLS and HPO reduces potential bias by standardizing variable definitions and removing the need for feature selection in favor of a hypothesis-free approach.

**Figure 1:**
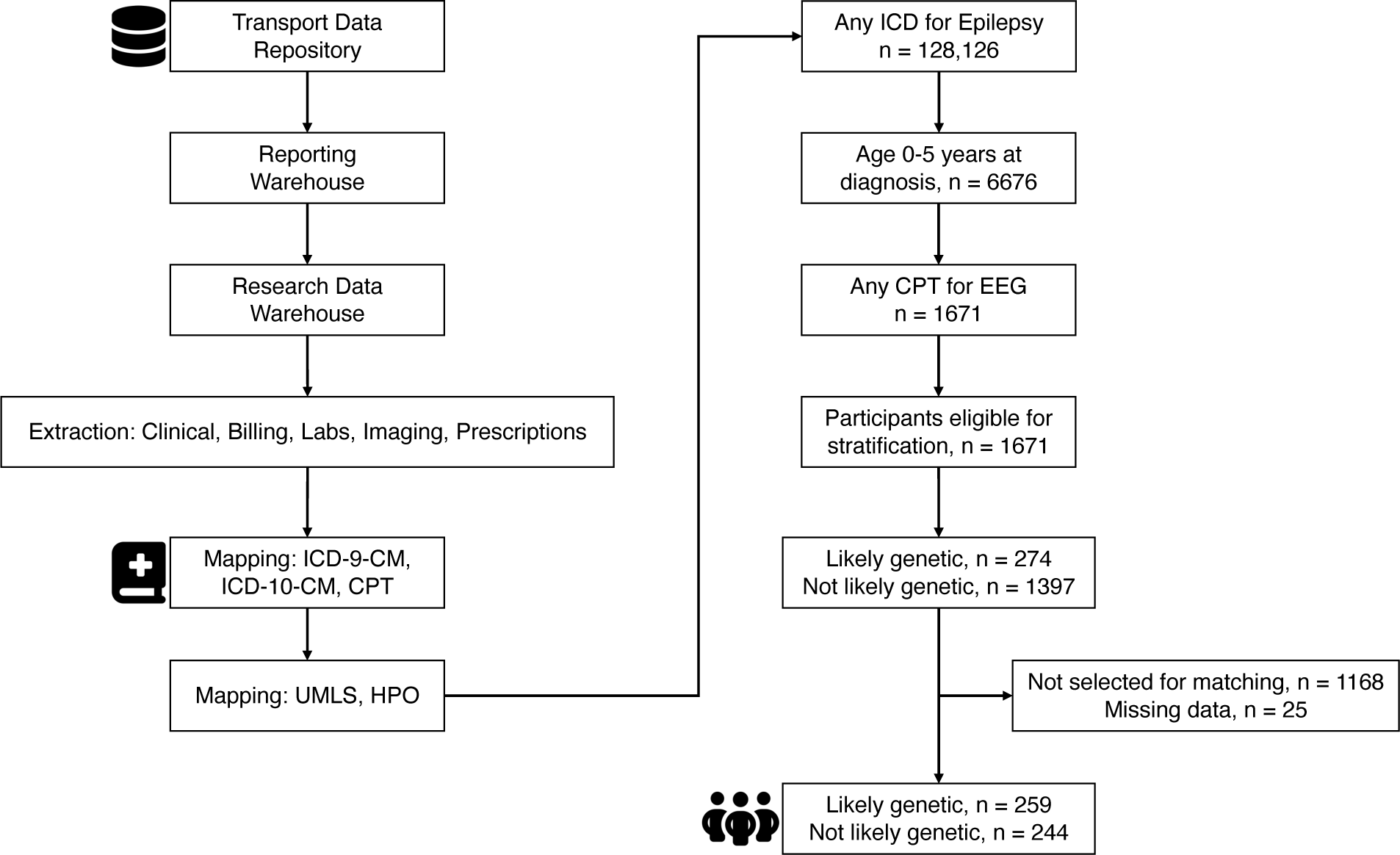
Flow diagram of datasets and processes used in the study. Abbreviations: CPT – Current Procedural Terminology; EEG – electroencephalography; EHR – electronic health record; HPO – Human Phenotype Ontology; ICD-9/10-CM – International Classification of Disease, Clinical Modification; UMLS – Unified Medical Language System.

After stratification, 25 individuals were removed due to missing data in encounter-date annotations which could not be confirmed as missing at random. No missing data imputation was done. Quantitative variables included age at the encounter, age at the last follow-up, and age at diagnosis. For longitudinal analyses, we grouped these according to the age ranges used by the ILAE Task Force on Nosology and Definitions: 0-2 years (neonatal/infantile), 2-12 years (childhood), 12-18 years (juvenile), and >18 years (adult).^19^

This study is reported according to the STROBE-RECORD extended checklist and meets all five CODE-EHR minimum best-practice framework standards for using structured healthcare data in clinical research.^20^

### Statistical analysis

This study was conducted in the R programming language, version 4.1.0, with RStudio, version 1.4.1106. We used two-sided Fisher’s exact or t-tests to test for association between categorical variables and genetic aetiology. The threshold for statistical significance was set to α = 0·05. P-values were adjusted for multiple testing with Bonferroni’s correction for whole-phenome analyses (i.e., association testing across all UMLS concepts or all HPO terms), and corrected p-values (p_adj_) are reported where appropriate. The effect sizes of relative enrichment were provided as odds ratios with 95% confidence intervals.

### Ethics statement

This study was approved by the Institutional Review Board of the Cleveland Clinic, approval IDs #22-147 and #23-253. Informed consent was waived due to the retrospective study design. All concept associations were deidentified to ensure data privacy, and all data was processed and stored on secure infrastructure.

## Results

### Healthcare resource utilization is higher in individuals with likely genetic epilepsy syndromes, most notably during the transition from pediatric to adult care

Genetic epilepsies are likely to have different healthcare utilization patterns that have not yet been quantified in a controlled study. Here, we included participants with childhood-onset epilepsy, where individuals with genetic epilepsy syndromes were identified by natural language processing. The final study cohort consisted of 259 individuals with known or likely genetic epilepsy syndromes and 244 matched controls (Table 1). Their ICD-10 syndrome diagnoses are shown in Table S2. The mean length of follow-up was 8·18 years (median 7, SD 5·01, range 0·10 – 21·70) for a cumulative follow-up of 4115 person-years (Figure 2A), and each individual had an average of 19·20 encounters within the healthcare system (median 11, SD 21·10, range 1 – 144). The median age at the first and last encounter was 0·1 years and 7·9 years, respectively. Electronic health record extraction yielded a total of 188,295 annotations across 9,659 encounters, with a mean of 8·94 unique Unified Medical Language System (UMLS) concepts (SD 8·62, range 1 – 54) and 19·2 HPO terms (SD 21·1, range 1 – 144) per individual. Each annotation corresponded to one single diagnostic or procedural concept mapped from raw text in clinical notes, billing information, or diagnostic results.

**Figure 2.**
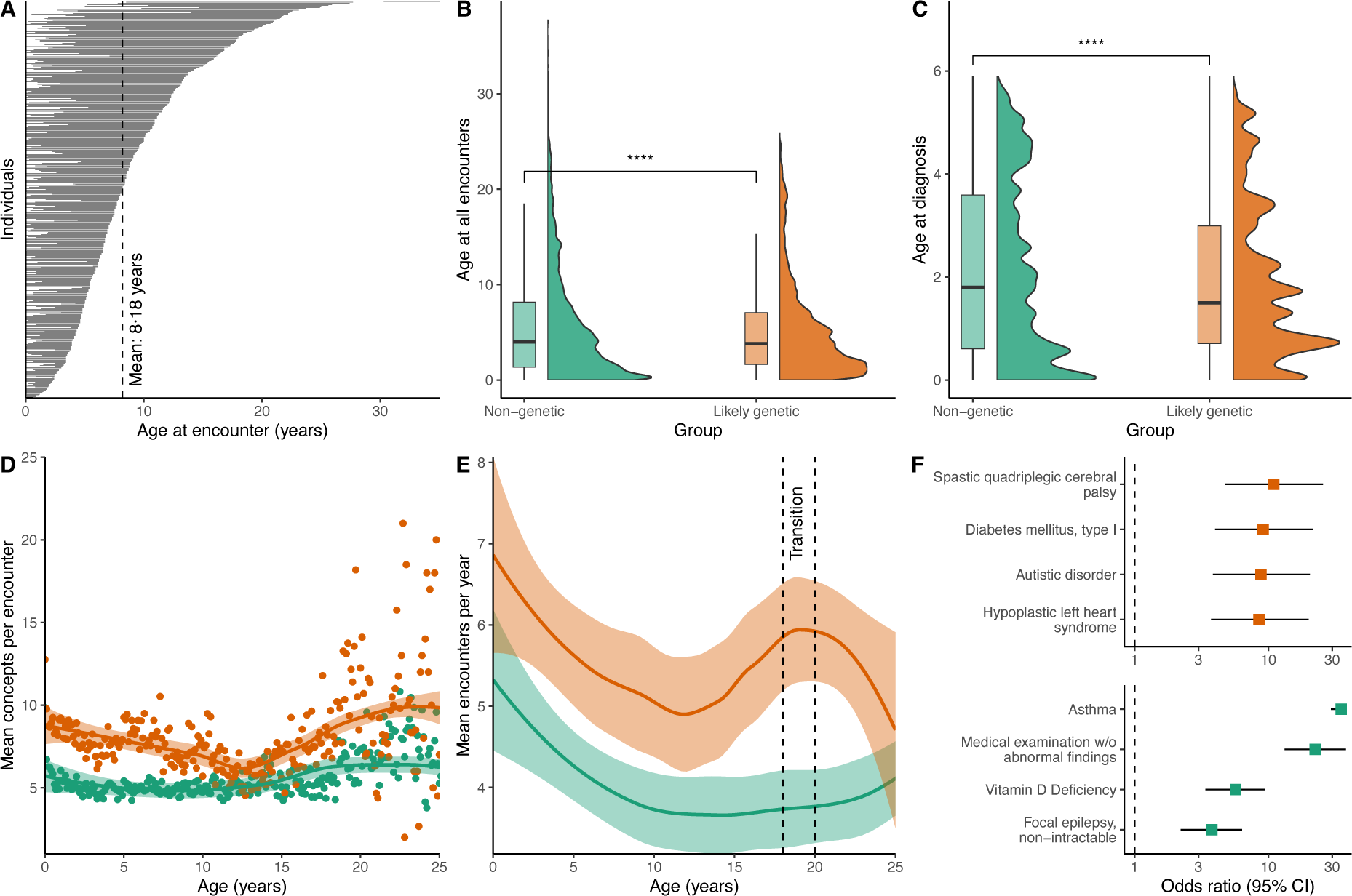
Length of follow-up, age distribution, and encounter distribution for the study cohort. **A:** Length of follow-up for each individual is shown as stacked horizontal lines, sorted by age at the last follow-up. Each line represents the length of EHR data available. **B:** Violin and boxplot of age at all encounters for individuals with likely genetic and non-genetic epilepsy syndromes. ****p < 0·0001. **C:** Violin and boxplot of age at diagnosis (fulfillment of eligibility criteria) for individuals with likely genetic and non-genetic epilepsy syndromes. ****p < 0·0001. **D:** Mean number of UMLS concepts per encounter for each group. Each dot is the mean number of monthly concepts per group. **E:** Mean number of annual encounters per year for each group. The line corresponds to the smooth conditional mean, with the shaded area being the standard error of the mean. The dashed lines mark the largest relative difference in annual encounter frequency, the transition period from pediatric to adult care (ages 18 – 20 years). **F:** Top four UMLS concepts with the greatest enrichment in the transition period. Forest plot of concept enrichment during the transition period compared to before the transition period, sorted by highest odds ratio and shown separately for each group.

**Table 1.**
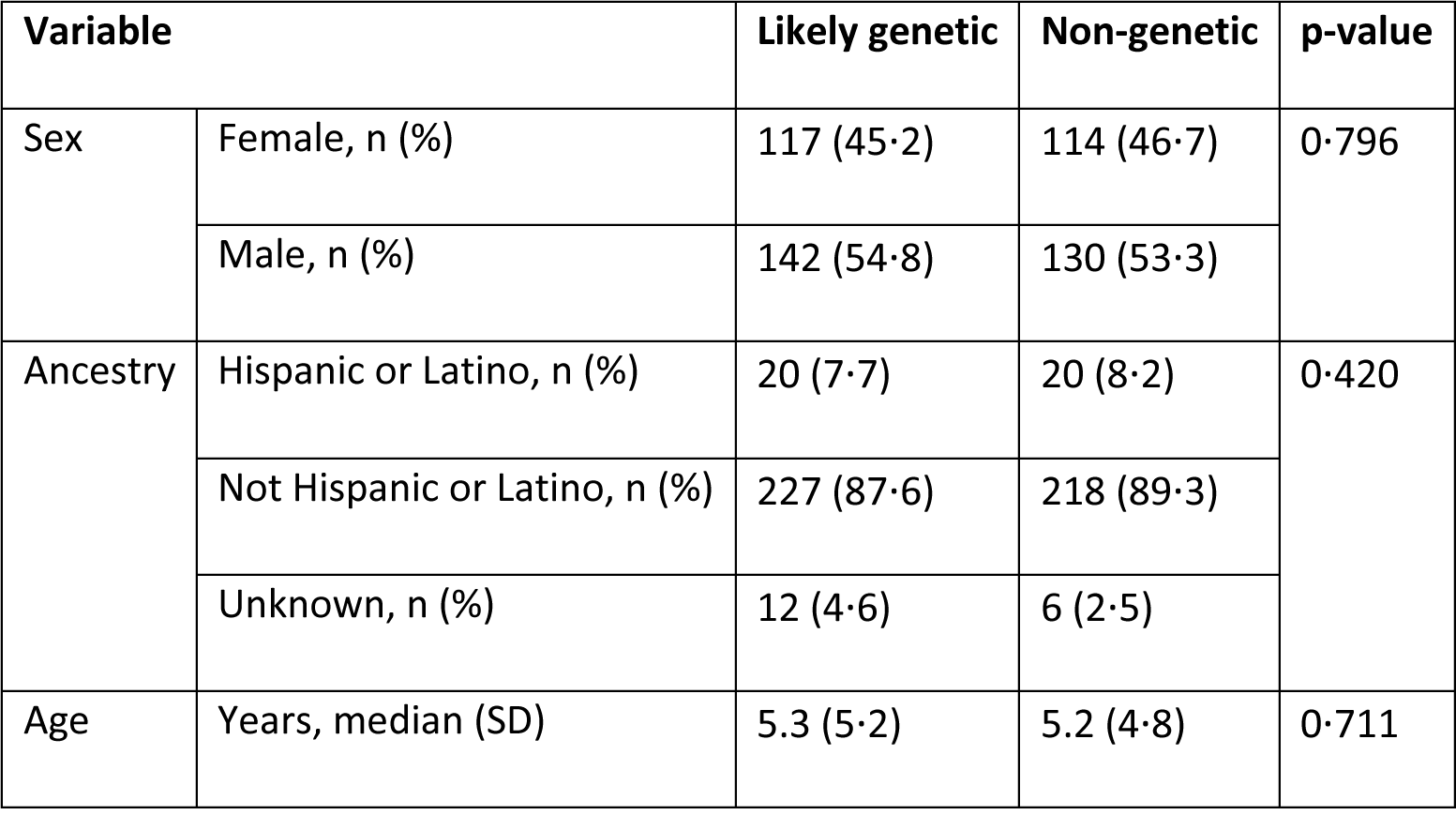
Demographic features of the study cohort.

| Variable |  | Likely genetic | Non-genetic | p-value |
| --- | --- | --- | --- | --- |
| Sex | Female, n (%) | 117 (45.2) | 114 (46.7) | 0.796 |
|  | Male, n (%) | 142 (54.8) | 130 (53.3) |  |
| Ancestry | Hispanic or Latino, n (%) | 20 (7.7) | 20 (8.2) | 0.420 |
|  | Not Hispanic or Latino, n (%) | 227 (87.6) | 218 (89.3) |  |
|  | Unknown, n (%) | 12 (4.6) | 6 (2.5) |  |
| Age | Years, median (SD) | 5.3 (5.2) | 5.2 (4.8) | 0.711 |

Individuals with likely genetic epilepsy syndromes were younger when they had any healthcare encounters (mean age 5·29 years vs. 5·83, two-sided t-test, p = 4·9×10^-7^, Figure 2B) and were younger when they were first diagnosed with epilepsy (age at ICD-10 G40.-, mean age 1·87 years vs. 2·09, two-sided t-test, p < 2·22×10^-16^, Figure 2C). Over the entire age range, individuals with likely genetic epilepsy received more annotations per encounter; mean concepts per encounter 8·13 (SD = 2·72) vs. 5·90 (SD = 2·71), two-sided t-test, p = 5·81×10^-22^ (Figure 2D), as a surrogate marker for phenotypic complexity or healthcare utilization. Out of the 354/503 (70%) of individuals admitted to the emergency department at least once, likely genetic individuals were admitted significantly more often; mean admissions 20·00 (SD = 16·30) vs. 12·30 (SD = 9·40), two-sided t-test, p = 1·06×10^-46^. Likewise, out of the 258/503 (51%) of individuals admitted to the inpatient service at least once, likely genetic individuals were significantly more likely to be admitted more often; mean admissions 12·40 (SD = 15·30) vs. 8·55 (SD = 8·78), two-sided t-test, p = 0·009.

Healthcare resource utilization may vary over time, and individuals with genetic epilepsy syndromes are known to require multidisciplinary care during the transition from pediatric to adult care.^21^ Indeed, the largest relative increase in annual encounters compared to controls was seen at ages 18 – 20 years; mean annual encounters 6·17 (SD = 3·26) vs. 3·63 (SD = 2·63), two-sided t-test, p = 3·3×10^-7^ (Figure 2E). Compared with encounters before transition, encounters in likely genetic individuals during the transition were enriched for cerebral palsy, autistic disorder, or severe somatic comorbidities. Encounters of non-genetic individuals were enriched for asthma, medical examinations without abnormal findings, or non-intractable epilepsy (Figure 2F).

### Individuals with likely genetic epilepsy syndromes have a distinct spectrum of associated clinical features

Individuals with likely genetic epilepsy syndromes may have distinct clinical features compared to controls with non-genetic epilepsy. We, therefore, extracted 188,295 annotations across 9,659 encounters from the EHR and established cross-sectional phenotypes by comparing the presence or absence of any of the >900,000 UMLS concepts and >13,000 HPO terms, with each hypothesis corrected for multiple testing. We report adjusted p-values (p_adj_) throughout this section. Test statistics showed only minimal p-value inflation (λ = 1 · 17, Figure 3A). UMLS concepts were used to reflect general diagnostics, as billing and procedural information may not directly map to phenotypic features represented in the HPO. Likely genetic individuals were enriched for UMLS concepts including chromosomal anomalies, intractable generalized epilepsy syndromes, and intellectual disability (Figure 3B). We used HPO terms to complement UMLS concepts for more detailed analyses across the entire clinical spectrum.

**Figure 3.**
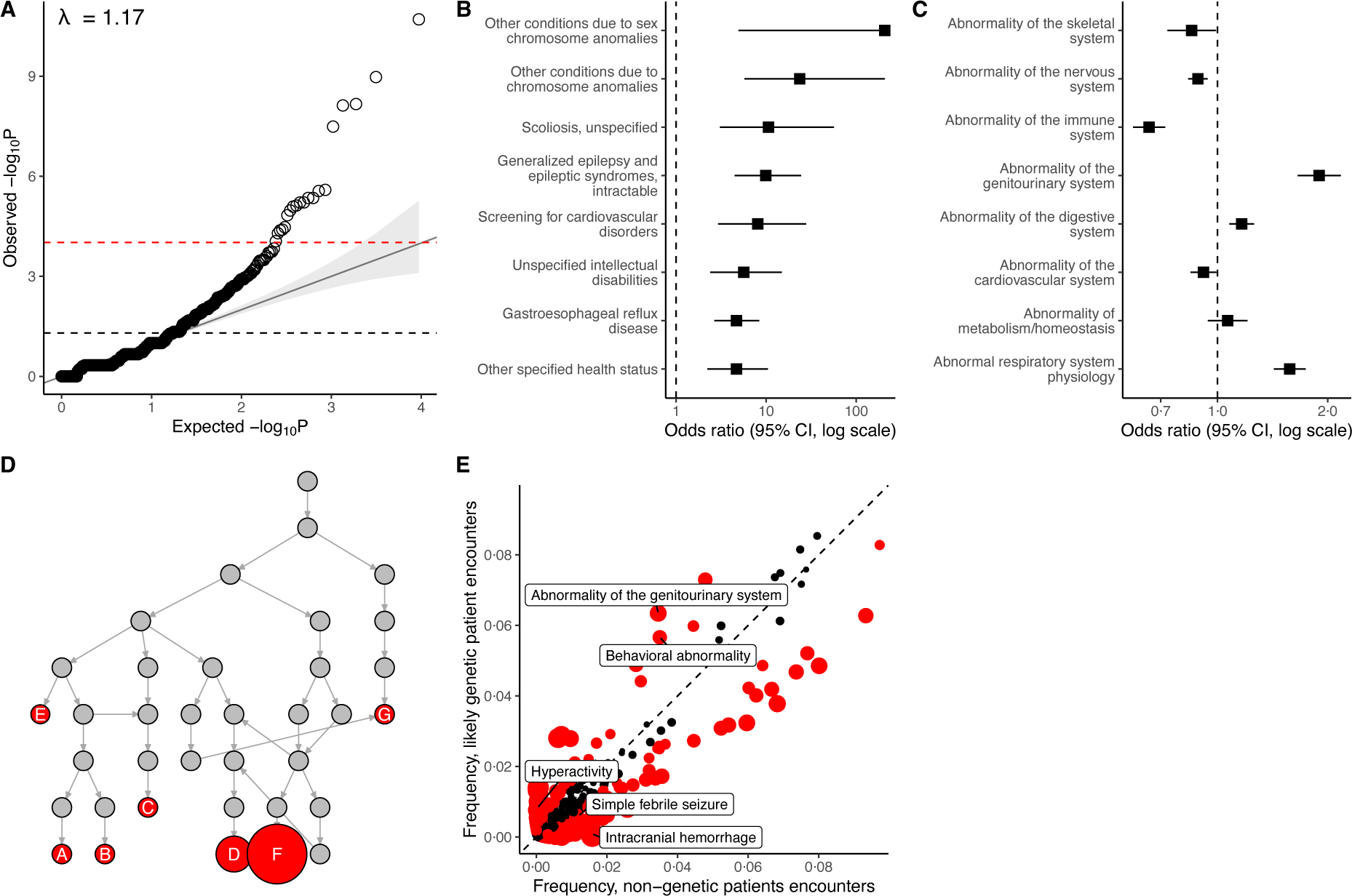
Cross-sectional analysis of clinical features associated with likely genetic epilepsy syndromes. **A:** Quantile-quantile (QQ) plot of the -log_10_ scaled nominal observed vs. expected p-value distribution for all tested hypotheses (UMLS concept association), showing minimal p-value inflation (λ = 1 · 17). The nominal significance threshold (α = 0 · 05) and Bonferroni-corrected significance threshold (α = 9 · 67*x*10^!”^) are shown as dashed lines. **B:** Forest plot of the top ten UMLS concepts most enriched in individuals with likely genetic epilepsy syndromes, sorted by Odds ratio. **C:** Forest plot of system-level HPO terms that are children of phenotypic abnormality (HP:0000118). **D:** Visualization of the subgraph rooted in an abnormality of the genitourinary system (HP:0000119). Nodes shown in red are terms that are independently significantly associated with individuals with likely genetic epilepsy and are labelled by the term they represent: A - ectopic kidney (HP:0000086), B - polycystic kidney dysplasia (HP:0000113), C - chronic kidney disease (HP:0012622), D - penile hypospadias (HP:0003244), E - abnormality of the ureter (HP:0000069), F - cryptorchidism (HP:0000028), G - urinary incontinence (HP:0000020). **E:** Relative frequency of HPO terms in encounters for individuals with likely genetic epilepsy syndromes versus those with non-genetic epilepsy syndromes. Each dot corresponds to a single term and is coloured red if significant.

We grouped annotations by system-level terms and noted that likely genetic individuals were enriched for abnormalities of the genitourinary system, a novel finding with a moderate effect size (HP:0000119; OR 1·91, 95% CI: 1·66 – 2·19, p_adj_ = 2·39×10^-19^, Figure 3C). Several clinical features contributed to this signal and were independently associated with likely genetic individuals: cryptorchidism (HP:0000028, p_adj_ = 2·62×10^-25^), penile hypospadias (HP:0003244, p_adj_ = 1·67×10^-15^), chronic kidney disease (HP:0012622, p_adj_ = 1·10×10^-7^), and others (Figure 3D). More fine-grained phenotypic representations are shown in Figure 3E, where we found likely genetic individuals to be enriched for behavioural abnormality (HP:0000708, OR 1·66, 95% CI: 1·45 – 1·90, p_adj_ = 5·64×10^-11^), including hyperactivity (HP:0000752, OR 24·71, 95% CI: 8·23 – 121·32, p_adj_ = 7·97×10^-18^), but depleted for simple febrile seizures (HP:0002373, OR 0·49, 95% CI: 0·45 – 0·70, p_adj_ = 2·97×10^-3^) and cerebral haemorrhage (HP:0001342, OR 0·02, 95% CI: 0·01 – 0·06, p_adj_ = 5·20×10^-46^), among others.

### Longitudinal analysis from childhood to adolescence reveals age-dependent patterns in clinical features and medical treatment

Genetic epilepsy syndromes are not static but represent dynamic entities with age-dependent clinical features. Identifying the timepoints where actionable phenotypes occur can inform diagnostic surveillance and clinical management. We, therefore, examined associated clinical features across age groups from infancy (0-2 years), childhood (2-12 years), youth (12-18 years), to adulthood (>18 years). Likely genetic individuals were significantly more likely to have recurrent infections (HP:0002719, OR 59·14, 95% CI: 10·43 – 2325·34, p_adj_ = 2·66×10^-17^), feeding difficulties (HP:0011968, OR 2·62, 95% CI: 2·15 – 3·23, p_adj_ = 1·23×10^-22^), constipation (HP:0002019, OR 2·92, 95% CI: 2·31 – 3·73, p_adj_ = 1·87×10^-21^), or dehydration (HP:0001944, OR 3·37, 95% CI: 2·31 – 5·08, p_adj_ = 3·00×10^-21^) in childhood (Figure 4A). Conversely, neonatal or infantile acquired causes of epilepsy were more likely in the non-genetic group, including cerebral haemorrhage (HP:0001342, OR 0·01, 95% CI: 0·01 – 0·02, p_adj_ = 1·64×10^-183^) and meningitis (HP:0001287, OR 0·00, 95% CI: 0·00 – 0·07, p_adj_ = 2·21×10^-10^). Interestingly, we found a strong signal for renal insufficiency in neonates and infants (HP:0000083, OR 43·50, 95% CI: 25·29 – 81·90, p_adj_ = 4·07×10^-170^), and osteoporosis in adults (HP:0000939, OR Inf, 95% CI: 3·73 – Inf, p_adj_ = 5·14×10^-3^) with known or likely genetic epilepsy syndromes. We included four common childhood comorbidities that were not expected to be enriched in cases (hyperglycemia, parasomnia, otitis media, and allergic rhinitis) as controls. Across the age range, none of these features were enriched in cases.

**Figure 4.**
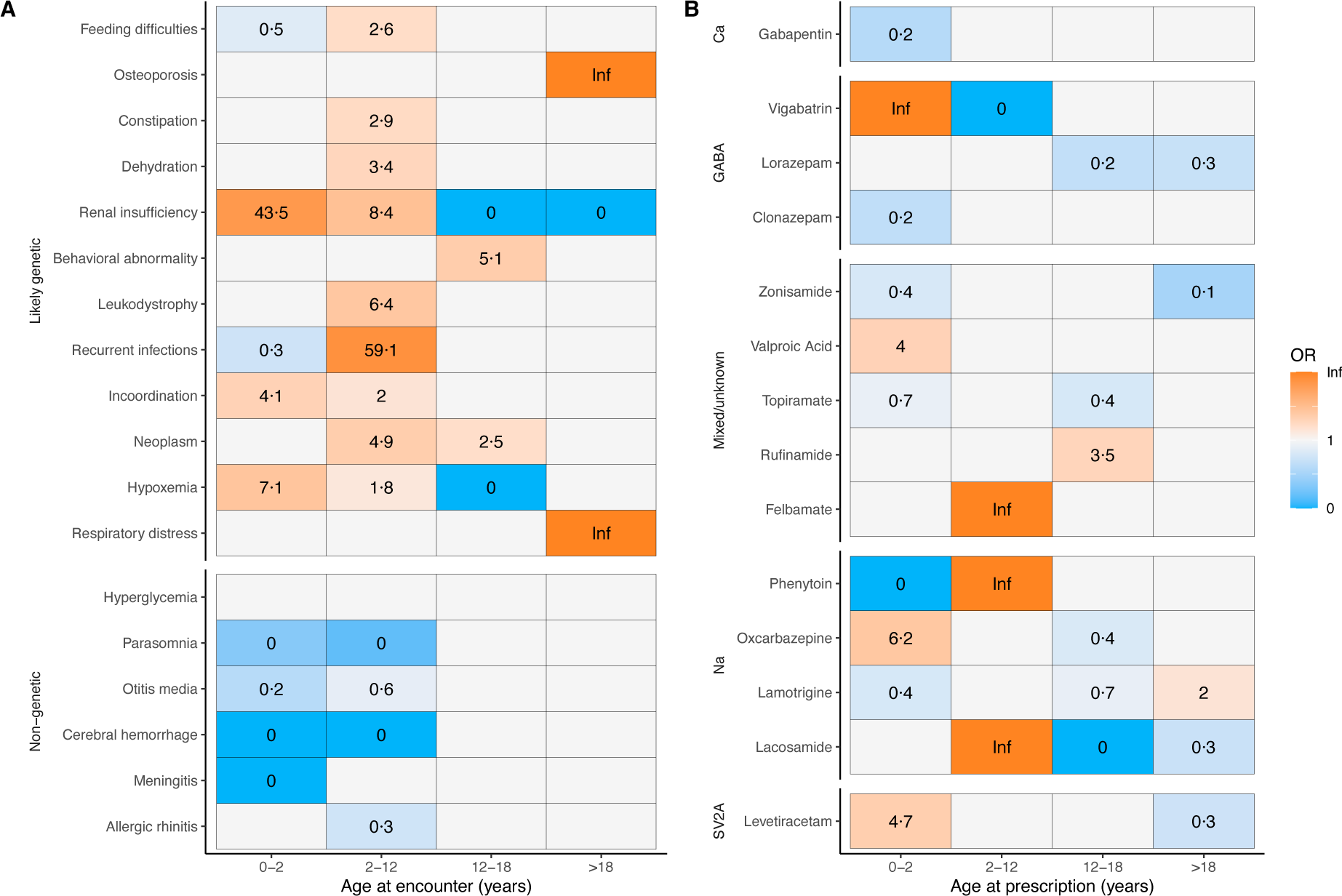
Longitudinal analysis of clinical features associated with likely genetic epilepsy syndromes. **A:** Heatmap of clinical features over the age ranges, binned by neonatal/infantile (0-2 years), childhood (2-12 years), juvenile (12-18 years), and adulthood (>18 years). Relative enrichment (odds ratio) of features between individuals with likely genetic epilepsy syndromes and those with non-genetic epilepsy syndromes is shown as labels. Blank tiles correspond to non-significant associations. Terms were grouped via hierarchical clustering of similar trajectories. **B:** Heatmap of anti-seizure medication (ASMs) prescription patterns, grouped by putative main mechanism of action. ASMs are shown if they had any significant group-level associations and were prescribed to at least 1% of the study cohort.

Likewise, we hypothesized that the treatment rationale of genetic epilepsy changes over the age range. Data on 15,003 prescriptions were available for 365/503 (73%) of the study cohort. Individuals with likely genetic epilepsy syndromes were more likely to receive long-term drug therapy (UMLS:C2911188, OR 4·32, 95% CI: 2·47 – 7·70, p_adj_ = 1·52×10^-7^) and received significantly more prior and concurrent anti-seizure medications (ASM); mean unique ASMs per person 4·47 (SD = 2·90) vs. 3·19 (SD = 2·24), two-sided t-test, p_adj_ = 3·04×10^-6^. Importantly, they received more prescriptions for rescue medication (benzodiazepines); mean prescriptions per person 11·90 (SD = 14·70) vs. 8·14 (SD = 13·40), two-sided t-test, p_adj_ = 0·048. Likewise, prescription patterns for ASM differed between the two groups and changed across age intervals. Individuals with likely genetic epilepsy syndromes received first-line ASMs (levetiracetam, valproic acid) earlier and broad-spectrum ASMs (phenytoin, lacosamide) later in life. Also, they were more likely to be exposed to syndrome-specific ASMs with potentially severe side effects (vigabatrin, felbamate, rufinamide) (Figure 4B).

### Data-driven identification of likely genetic individuals reveals unexpected and underrecognized aetiologies beyond common genetic epilepsy syndromes

We validated our key findings with manual chart review for 45 cases, focusing on individuals with the potentially novel phenotypic associations outlined above: renal insufficiency in neonates and infants, osteoporosis in adulthood, and genitourinary abnormalities (Table 2). Of these, 30/45 cases (66%) had a confirmed genetic diagnosis (not considering variants of unknown significance), 4/45 (8·9%) had genetic testing in progress at the last follow-up, 3/45 (6·6%) had negative results on genetic testing, 1/45 (2%) declined genetic testing, and the rest were lost to follow-up. Neonatal and infantile renal insufficiency or genitourinary abnormalities were primarily observed in rare congenital multisystem disorders (e.g., Kabuki syndrome, Warburg Micro syndrome, DiGeorge syndrome, or Cornelia de Lange syndrome) and microdeletion or duplication syndromes (e.g., chromosome 15q11-q13 duplication, Prader-Willi syndrome). In all cases, osteoporosis was confirmed by a DEXA scan and was found in childhood hypophosphatasia, combined oxidative phosphorylation deficiency, and Dravet syndrome. Canonical genetic epilepsy syndromes (e.g., tuberous sclerosis complex 1, *CDKL5*-related developmental and epileptic encephalopathy 2, or ion channel disorders) comprised only the minority of cases (Table 2).

**Table 2.**
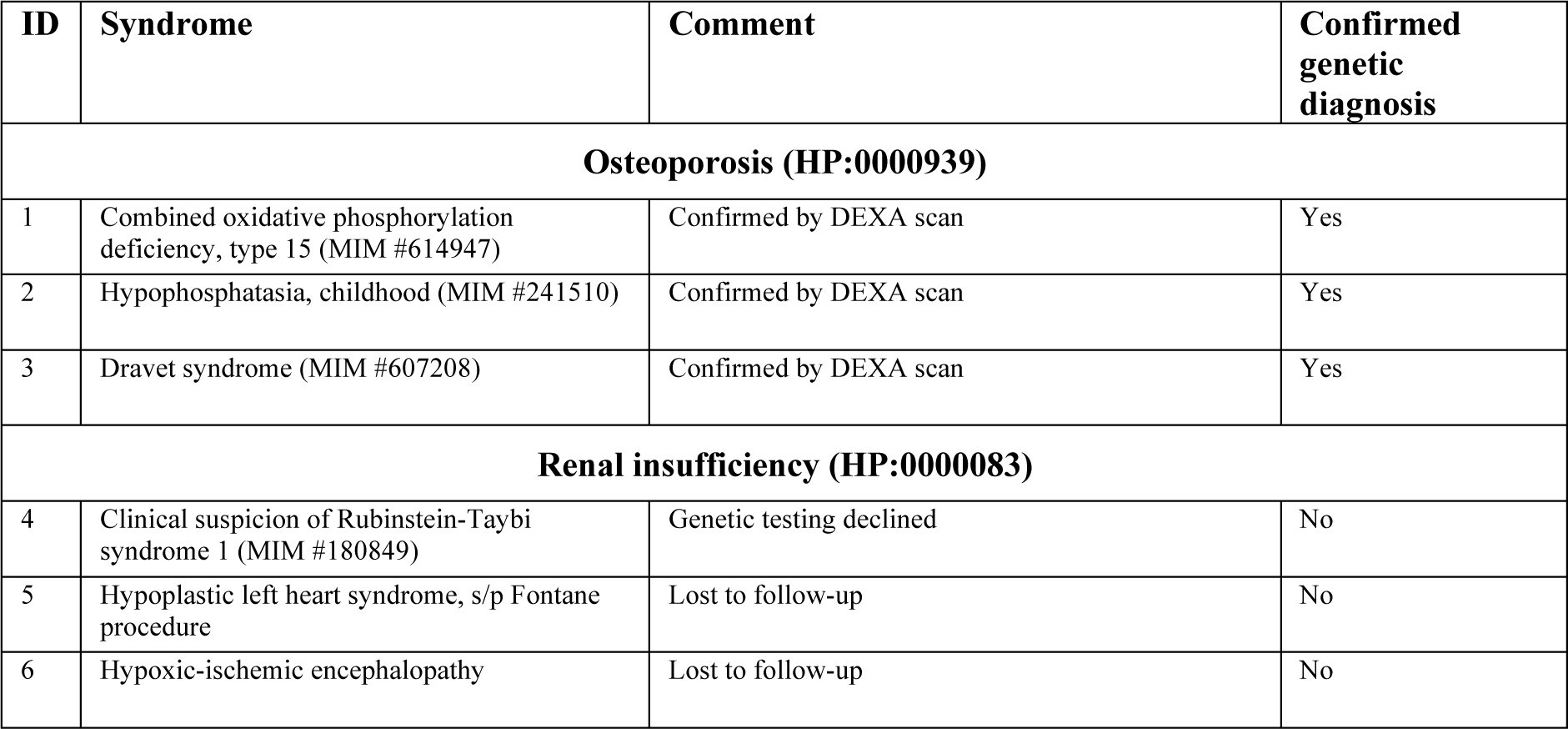
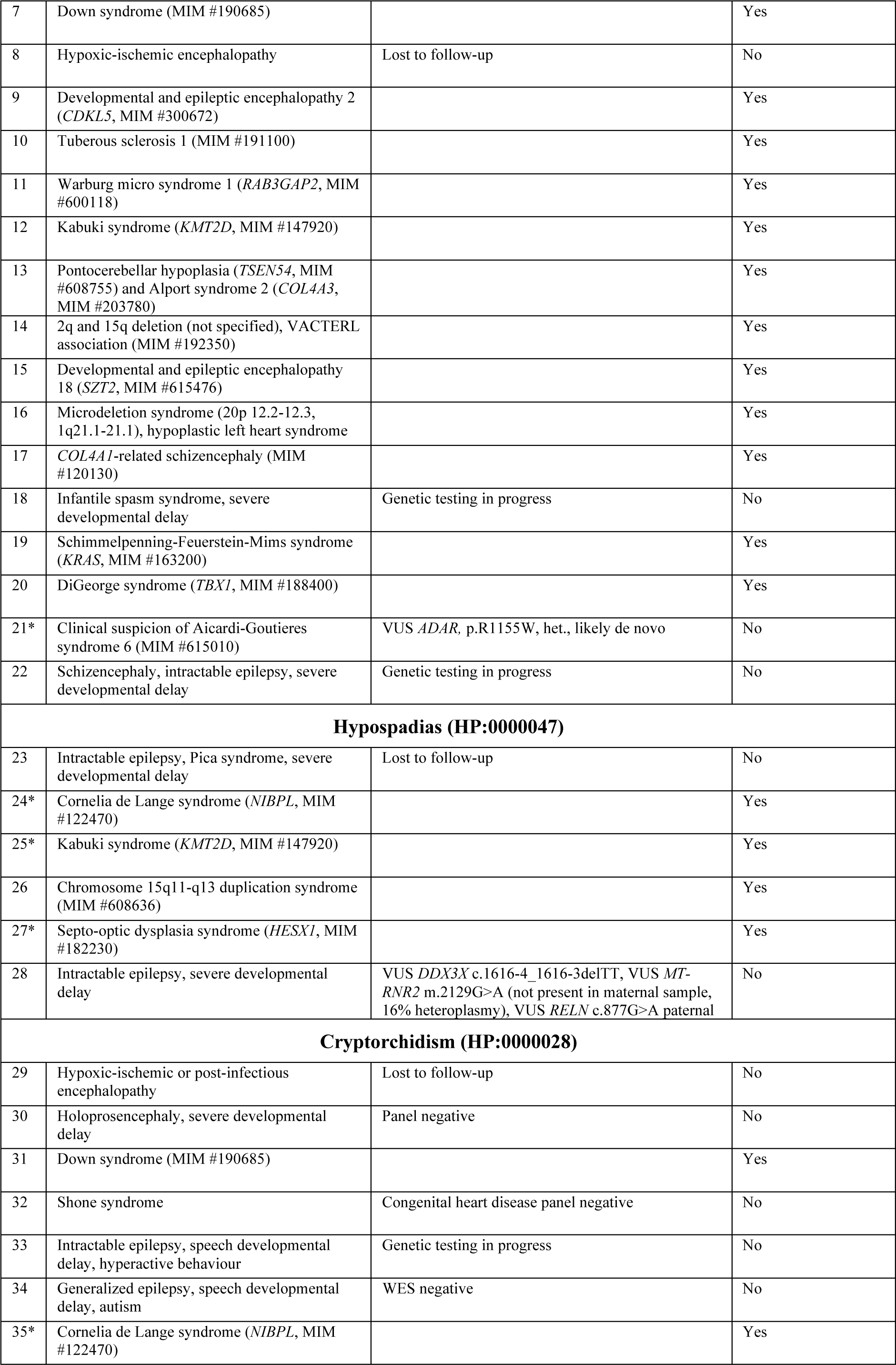
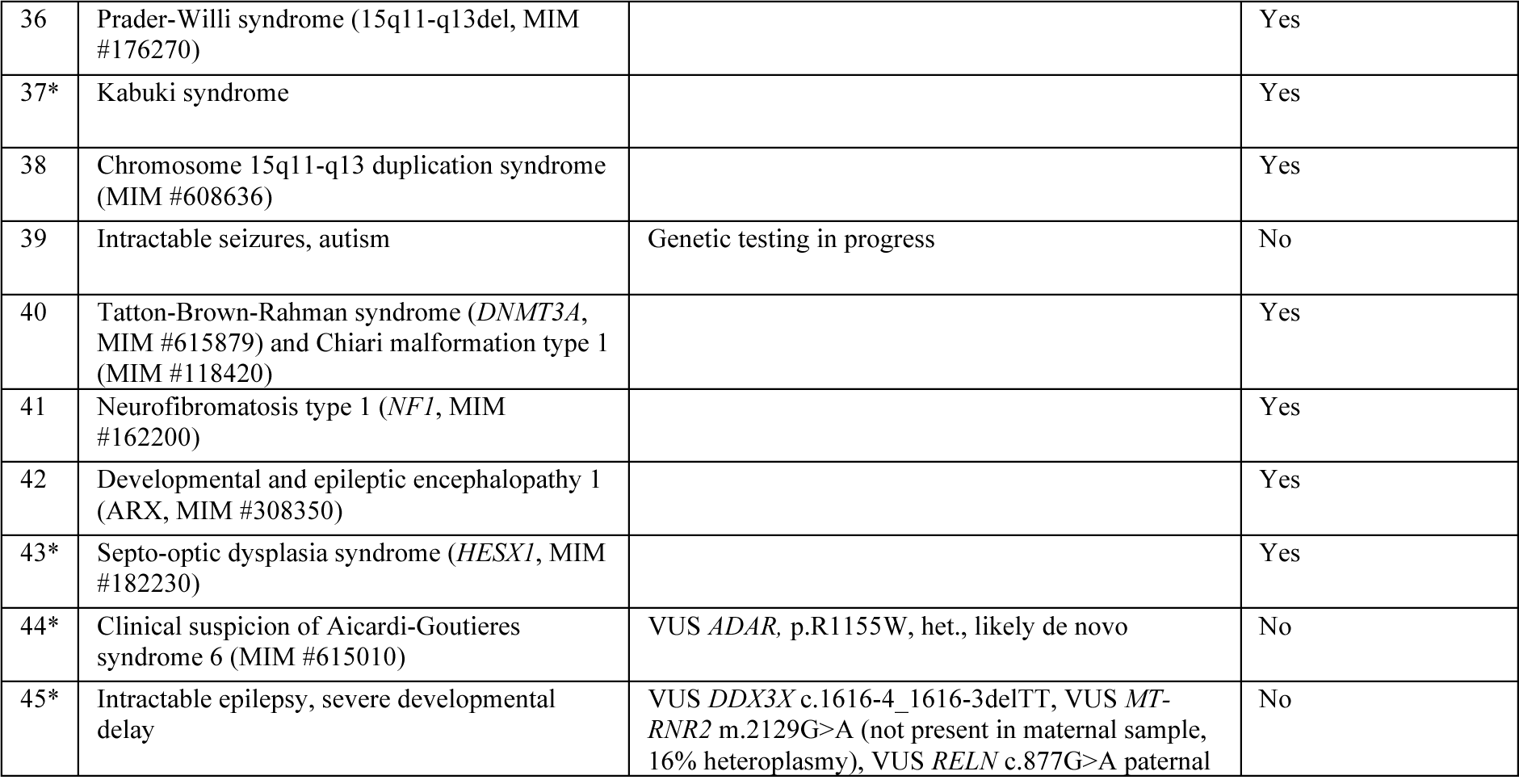
Results of manual chart view to confirm key novel findings. Each row corresponds to one study participant, grouped by key phenotypic features (HPO terms) that were found to be associated with a likely genetic diagnosis on cross-sectional and longitudinal analysis. Due to phenotypic overlap, some individuals are represented in several groups and are marked with (*). A confirmed genetic diagnosis is indicated by presence of a disease-causing variant on chart review, not counting variants of unknown significance (VUS), and is reported here to demonstrate the performance of our phenotyping algorithm. Abbreviations: DEXA – dual-energy x-ray absorptiometry; MIM – Mendelian Inheritance in Man; VUS – variant of unknown significance; WES – whole-exome sequencing.

| ID | Syndrome | Comment | Confirmed genetic diagnosis |
| --- | --- | --- | --- |
| **Osteoporosis (HP:0000939)** |  |  |  |
| 1 | Combined oxidative phosphorylation deficiency, type 15 (MIM #614947) | Confirmed by DEXA scan | Yes |
| 2 | Hypophosphatasia, childhood (MIM #241510) | Confirmed by DEXA scan | Yes |
| 3 | Dravet syndrome (MIM #607208) | Confirmed by DEXA scan | Yes |
| **Renal insufficiency (HP:0000083)** |  |  |  |
| 4 | Clinical suspicion of Rubinstein-Taybi syndrome 1 (MIM #180849) | Genetic testing declined | No |
| 5 | Hypoplastic left heart syndrome, s/p Fontane procedure | Lost to follow-up | No |
| 6 | Hypoxic-ischemic encephalopathy | Lost to follow-up | No |
| 7 | Down syndrome (MIM #190685) |  | Yes |
| 8 | Hypoxic-ischemic encephalopathy | Lost to follow-up | No |
| 9 | Developmental and epileptic encephalopathy 2 (*CDKL5*, MIM #300672) |  | Yes |
| 10 | Tuberous sclerosis 1 (MIM #191100) |  | Yes |
| 11 | Warburg micro syndrome 1 (*RAB3GAP2*, MIM #600118) |  | Yes |
| 12 | Kabuki syndrome (*KMT2D*, MIM #147920) |  | Yes |
| 13 | Pontocerebellar hypoplasia (*TSEN54*, MIM #608755) and Alport syndrome 2 (*COL4A3*, MIM #203780) |  | Yes |
| 14 | 2q and 15q deletion (not specified), VACTERL association (MIM #192350) |  | Yes |
| 15 | Developmental and epileptic encephalopathy 18 (*SZT2*, MIM #615476) |  | Yes |
| 16 | Microdeletion syndrome (20p 12.2-12.3, 1q21.1-21.1), hypoplastic left heart syndrome |  | Yes |
| 17 | *COL4A1*-related schizencephaly (MIM #120130) |  | Yes |
| 18 | Infantile spasm syndrome, severe developmental delay | Genetic testing in progress | No |
| 19 | Schimmelpenning-Feuerstein-Mims syndrome (*KRAS*, MIM #163200) |  | Yes |
| 20 | DiGeorge syndrome (*TBX1*, MIM #188400) |  | Yes |
| 21* | Clinical suspicion of Aicardi-Goutieres syndrome 6 (MIM #615010) | VUS *ADAR*, p.R1155W, het., likely de novo | No |
| 22 | Schizencephaly, intractable epilepsy, severe developmental delay | Genetic testing in progress | No |
| **Hypospadias (HP:0000047)** |  |  |  |
| 23 | Intractable epilepsy, Pica syndrome, severe developmental delay | Lost to follow-up | No |
| 24* | Cornelia de Lange syndrome (*NIBPL*, MIM #122470) |  | Yes |
| 25* | Kabuki syndrome (*KMT2D*, MIM #147920) |  | Yes |
| 26 | Chromosome 15q11-q13 duplication syndrome (MIM #608636) |  | Yes |
| 27* | Septo-optic dysplasia syndrome (*HESX1*, MIM #182230) |  | Yes |
| 28 | Intractable epilepsy, severe developmental delay | VUS *DDX3X* c.1616-4_1616-3delTT, VUS *MT-RNR2* m.2129G>A (not present in maternal sample, 16% heteroplasmy), VUS *RELN* c.877G>A paternal | No |
| **Cryptorchidism (HP:0000028)** |  |  |  |
| 29 | Hypoxic-ischemic or post-infectious encephalopathy | Lost to follow-up | No |
| 30 | Holoprosencephaly, severe developmental delay | Panel negative | No |
| 31 | Down syndrome (MIM #190685) |  | Yes |
| 32 | Shone syndrome | Congenital heart disease panel negative | No |
| 33 | Intractable epilepsy, speech developmental delay, hyperactive behaviour | Genetic testing in progress | No |
| 34 | Generalized epilepsy, speech developmental delay, autism | WES negative | No |
| 35* | Cornelia de Lange syndrome (*NIBPL*, MIM #122470) |  | Yes |
| 36 | Prader-Willi syndrome (15q11-q13del, MIM #176270) |  | Yes |
| 37* | Kabuki syndrome |  | Yes |
| 38 | Chromosome 15q11-q13 duplication syndrome (MIM #608636) |  | Yes |
| 39 | Intractable seizures, autism | Genetic testing in progress | No |
| 40 | Tatton-Brown-Rahman syndrome (*DNMT3A*, MIM #615879) and Chiari malformation type 1 (MIM #118420) |  | Yes |
| 41 | Neurofibromatosis type 1 (*NF1*, MIM #162200) |  | Yes |
| 42 | Developmental and epileptic encephalopathy 1 (ARX, MIM #308350) |  | Yes |
| 43* | Septo-optic dysplasia syndrome (*HESX1*, MIM #182230) |  | Yes |
| 44* | Clinical suspicion of Aicardi-Goutieres syndrome 6 (MIM #615010) | VUS *ADAR*, p.R1155W, het., likely de novo | No |
| 45* | Intractable epilepsy, severe developmental delay | VUS *DDX3X* c.1616-4_1616-3delTT, VUS *MT-RNR2* m.2129G>A (not present in maternal sample, 16% heteroplasmy), VUS *RELN* c.877G>A paternal | No |

## Discussion

Healthcare resource utilization and disease burden in individuals with genetic epilepsy syndromes are not well-understood, as these syndromes are individually rare. Previous studies have attempted to address this problem by observing direct costs or quality of life from insurance claims and online surveys.^22^ Here, we instead utilized natural language processing and deep computational phenotyping across a large healthcare system to identify a longitudinal cohort of individuals with childhood-onset likely genetic epilepsy syndromes and matched controls with non-genetic epilepsy. We found several markers of increased healthcare resource utilization. Individuals with likely genetic epilepsy syndromes were more likely to be admitted to inpatient services or the emergency department. They had more frequent encounters with the healthcare system and more diagnoses per encounter. Importantly, they were seen significantly more often during the transition from pediatric to adult care, likely because of more severe comorbidities. Transition is a critical period that requires multidisciplinary care teams.^21^ This study provides objective evidence to support the need for transition care, which was previously limited.^21^

Finding clinical features associated with genetic epilepsy syndromes improves patient selection and cost-effectiveness for genetic testing by increasing the pre-test probability of a positive finding.^14^ Previous studies have demonstrated how deep longitudinal data from healthcare systems can be leveraged to characterize monogenic syndromes.^7–9^ Here, we validated previous findings, including independent statistical support for several known predictors: intractable seizures, behavioral abnormalities, autism, developmental delay, intellectual disability, abnormalities of movement (including ataxia), pharmacoresistance (long-term drug therapy), and others. Conversely, we found individuals with probable causes of acquired epilepsy (e.g., cerebral hemorrhage, meningitis) less likely to have a genetic diagnosis. These factors have been described in studies of clinical sequencing yield, which are reflected in current practice guidelines that recommend genetic testing, preferably whole-exome sequencing, in any individual with seizures and intellectual disability.^14,15^

Our data-driven whole-phenome approach identified individuals with syndromes that commonly present with seizures, but which are not traditionally considered epilepsy syndromes. These include rare congenital multisystem disorders and chromosomal disorders, which have received less attention when compared to the aetiology-specific developmental and epileptic encephalopathies caused by ion channel or transporter disorders.^23^ In our study, these individuals contributed towards a novel signal for genitourinary abnormalities including congenital malformations. This clinical aspect can therefore be kept in mind for children with dysmorphic and chromosomal syndromes. Further, longitudinal phenotyping revealed markers of disease burden and age-specific general clinical features, e.g., a higher likelihood of feeding difficulties, dehydration, constipation, recurrent infections, or hypoxemia in childhood. These are clinical issues commonly seen in neurodevelopmental disorders.^24^ Likewise, data from medical prescriptions demonstrated group-level differences in disease burden and severity. Individuals with likely genetic epilepsy syndromes were more likely to receive long-term drug therapy, with more prescriptions for rescue medication and earlier exposure to broad-spectrum or syndrome-specific ASMs, in line with previous evidence of polypharmacy in this vulnerable group.^25^

This study leveraged >180,000 concept annotations across >4000 person-years, utilizing deep computational phenotyping and well-matched controls to provide statistical power for our analysis. The study site, an integrated Level 4 Adult and Paediatric Epilepsy Centre enabled us to achieve longer follow-up than previous studies, spanning the critical transition period. The cohort definition was based on gold-standard criteria, with orthogonal validation by another scoring system and manual chart review. Our hypothesis-free ontological reasoning approach was designed to minimize the effect of bias or unaccounted confounders.

However, this study only reports on statistical associations and cannot be used to establish causality between genetic syndromes and their comorbidities. We note that some of the associations, e.g., osteoporosis and renal insufficiency, may be secondary due to malnutrition, drug side effects, or multi-organ dysfunction. A potential risk of misclassification bias may be addressed by extending recent work on machine-learning-based patient identification.^26^ While this study was conducted in a large multi-center healthcare system, we were still limited to a US population sample. As demonstrated above, independent replication of findings and external validity across different healthcare systems remains a central challenge. Lastly, healthcare systems as data sources will always be subject to key limitations, including documentation quality variability, billing or procedural practice changes, and discontinuous healthcare usage.^27^

Future research directions may include deep computational phenotyping in clinical sequencing yield studies to power gene discovery and confirm the clinical utility of the identified statistical associations. Finally, an improved understanding of the longitudinal disease trajectories of these individuals will contribute towards both a timely diagnosis and syndrome-specific disease forecasting models.^28^

## Contributors

Supervision: DL. Methodology: CMB. Data Curation: CMB, AI, MSJ, AM. Data Validation: CMB, AI, MSJ, AM. Formal analysis: CMB. Writing – Original Draft: CMB. Writing – Review & Editing: AI, MSJ, AM, CL, EPK, AG, IN, DL. All authors have read and approved the final version of the manuscript.

## Data Availability

Deidentified individual participant data can be made available upon reasonable requests submitted to the corresponding author. The prerequisite for data sharing is a data transfer agreement approved by the legal departments and institutional review board of the requesting researcher. After proposal approval, data can be shared through a secure online platform. All code used for data analysis and visualization is available at https://github.com/christianbosselmann/UMLS-HPO.

## Research in Context

### Evidence before this study

Recent advances in natural language processing and electronic health record mining have enabled deep and longitudinal phenotyping of rare genetic epilepsy syndromes. We conducted a literature search using the PubMed database for articles published between 01/01/2010 and 01/03/2022 using the search terms (genetic) AND (epilepsy OR seizures OR seizure) AND (electronic health record OR electronic medical record). The 114 results identified by the custom PubMed search were filtered down to four papers describing computational phenotyping in genetic epilepsy syndromes. These four identified studies included previous work by Helbig and colleagues primarily involving single-gene or gene-family phenotypes in a pediatric cohort and a recent longitudinal analysis of a more general cohort by Ganesan et al.

### Added value of this study

Here, we present the first case-control study that uses deep computational phenotyping from electronic health records (EHR) to investigate individuals with childhood-onset epilepsy. Our novel natural language processing approach accurately stratified patients by the likelihood of an underlying genetic aetiology. Longitudinal phenotyping from EHR represents a rich data source that allowed us to analyze age-dependent patterns of healthcare resource utilization, medical treatment, and encounters with the healthcare system. The study setting, a comprehensive paediatric and adult epilepsy center, enabled us to achieve the longest mean follow-up compared to previous studies, for the first time including new insight on the critical transition stage from paediatric to adult neurological care. We found clinical features that are independently associated with a likely diagnosis of a genetic epilepsy syndrome, both robustly quantifying previously published data and highlighting several novel findings, such as genitourinary abnormalities linked to a spectrum of likely underrecognized and underdiagnosed congenital disorders.

### Implications of all the available evidence

Individuals with genetic epilepsy syndromes suffer from high unmet medical needs. Their healthcare resource utilization is higher than that of individuals with non-genetic epilepsy syndromes, especially during the transition from paediatric to adult care. Overall, they are affected by a severe disease burden from somatic and psychiatric comorbidities, as well as polypharmacy with anti-seizure medications. The clinical characteristics identified in this study will inform clinical surveillance and management. Finally, this data will help clinicians identify individuals that are suitable candidates for genetic testing, contributing towards cost-effective resource utilization for healthcare systems and a timely diagnosis for these often severely affected individuals.

## Abbreviations

ASM: anti-seizure medication
CPT: Current Procedural Terminology
DEE: developmental and epileptic encephalopathies
EEG: electroencephalography
EHR: electronic health records
HPO: human phenotype ontology
ICD: International Classification of Diseases
ILAE: International League Against Epilepsy
NLP: natural language processing
UMLS: Unified medical language system
QQ: quantile-quantile

## Acknowledgements

None.

