## Supplementary Material for "Healthcare utilization and clinical characteristics of genetic epilepsy syndromes: a longitudinal case-control study of electronic health records"

### Supplementary Figure 1: Additional validation with the PheIndex criteria

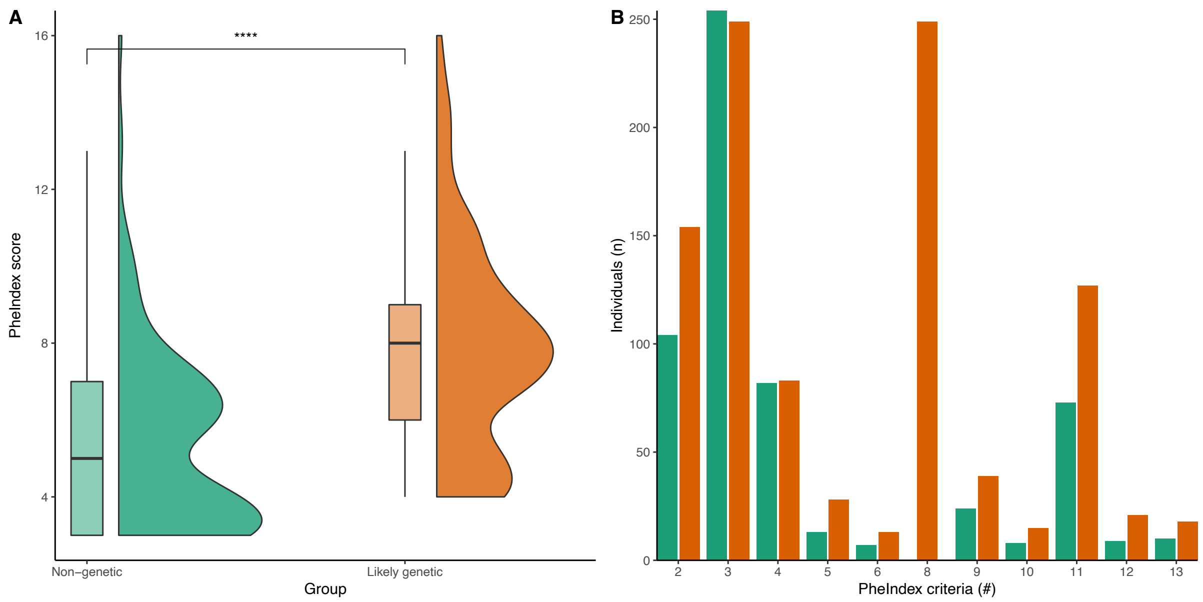

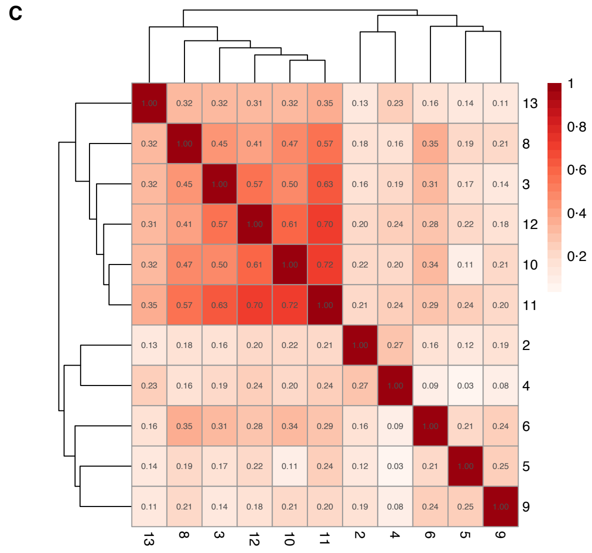

D

| Criterion | Description |
|---|---|
| 1 | Prolonged stay in the NICU |
| 2 | Prolonged or multiple hospitalizations |
| 3 | Visits or consults with multiple specialists |
| 4 | Multiple emergency room visits |
| 5 | Feeding support |
| 6 | Respiratory support |
| 7 | Imaging |
| 8 | Genetic diagnostic tests |
| 9 | Metabolic diagnostic tests |
| 10 | In−hospital death |
| 11 | Developmental delay |
| 12 | Diagnosis corresponding to metabolic diseases |
| 13 | Heart surgeries |

**Figure S1. PheIndex scores for individuals with likely genetic epilepsy syndromes and individuals with non-genetic epilepsy syndromes. A:** Likely genetic patients had a significantly higher average PheIndex score (7·84 SD 2·71 vs. 5·41 SD 2·41, two-sample t-test, $p = 9{\cdot}79 \times 10^{-24}$). **B:** Distribution of PheIndex criteria for both groups. Data for criterion 1 (prolonged stay in the neonatal intensive care unit, NICU) and criterion 7 (imaging) were not available. Note that criterion 3 is true for both groups (due to the study setting, a multidisciplinary epilepsy centre), and criterion 8 is true for cases only (due to the eligibility criteria including genetic testing). **C:** Heatmap of co-occurrence of PheIndex criteria in the study cohort. Developmental delay, in-hospital death, metabolic disease, consults with multiple specialists, and genetic diagnostic tests formed a single cluster. Labels show the pairwise frequency at which two criteria occur in the same individual. Hierarchical clustering on cosine similarity between sets of PheIndex criteria. **D:** List of PheIndex criteria.

### Supplementary Figure 2: Ontological representation and propagation

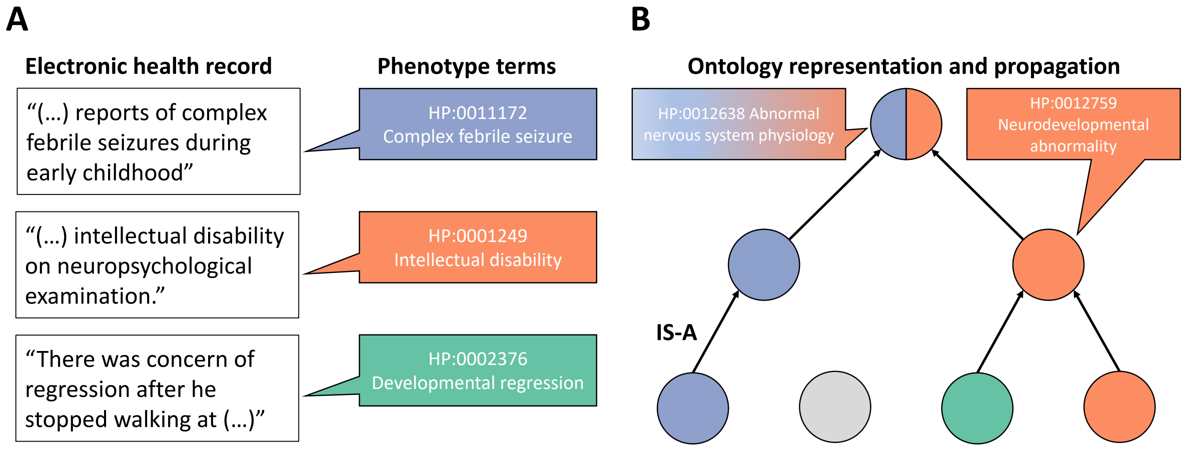

**Figure S2. A:** Clinical descriptions from provider notes, billing codes, and other text sources are extracted from the electronic health record (EHR) and mapped to standardized human phenotype ontology (HPO) terms. Each term represents a node in a directed acyclic graph, where edges between nodes are IS-A (parent-child) relationships. Terms are propagated along these edges, where each parent term is included in the set of terms until the root node (i.e., HP:0000001 All) for each original term is included in the set. **B:** Intuitively, this allows the data to reflect the hierarchical ordering of clinical findings, i.e., that terms like developmental regression and intellectual disability both fall under the broader category of neurodevelopmental abnormality, which in turn is an abnormality of nervous system physiology. Figure adapted from Boßelmann et al.[1]

### Supplementary Table 1: Natural language processing algorithm

Participants eligible for stratification were filtered for electronic health record (EHR) notes containing a set of prespecified epilepsy-related genes: *CACNA1A, CACNA1E, CDKL5, DEPDC5, GABRA1, GABRB2, GABRB3, GABRG2, GNAO1, GRIN1, GRIN2A, GRIN2B, GRIN2D, KCNA1, KCNA2, KCNB1, KCNQ2, KCNT1, MECP2, MTOR, NPRL2, NPRL3, PCDH19, PIK3CA, SCN1A, SCN2A, SCN3A, SCN8A, SLC13A5, SLC2A1, SLC6A1, STX1B, STXBP1, SYNGAP1, TSC1, TSC2*. These notes (n = 230) were then manually reviewed for sets of phrases that either confirmed or disproved that the individual had a pathogenic or likely pathogenic variant or variant of unknown significance (VUS) (Table S1). For each phrase, we assigned a score of negatively correlated (-1) or positively correlated (+1) with the presence of a genetic aetiology (Table S1). For each individual, phrases across all EHR notes were aggregated to calculate the final sum score. Note sections pertaining to family history or insurance approval requests were removed from analysis. Any individual with a sum score > 0 was considered to have a likely genetic aetiology.

| Negatively correlated phrases | Positively correlated phrases |
|---|---|
| 'sequencing {gene} negative' | 'confirmed {gene} mutation' |
| '{gene} negative mutation' | 'positive {gene}.* c.' |
| '{gene} mutation negative' | '{gene} gene mutation' |
| 'sequencing {gene} normal' | '{gene} genetic mutation' |
| 'genetic test.*{gene} normal' | 'mutation {gene} gene' |
| '{gene} negative' | 'mutation gene {gene}' |
| '{gene} negative' | 'mutation {gene} gene' |
| 'negative {gene}' | '{gene} gene sequencing identified mutation' |
| 'negative mutation analysis of {gene}' | '{gene} gene pathogenic mutation' |
| '{gene} test negative' | 'pathogenic {gene}.{0,10}mutation' |
| 'gene test {gene} neg ' | 'variants unknown significance {gene}' |
| 'negative mutation {gene}' | 'pathogenic variant {gene}' |
| '{gene} gene mutation negative' | 'variants uncertain significance detected {gene}' |
| '{gene} gene test negative' | 'remarkable {gene}' |
| 'No mutations {0,10} {gene}' | '{gene} mutation .{0,10} [0-9][ACG][<>]' |
| 'tested genetic.{0,10}{gene} neg ' | '{gene} Variant Transition Nucleotide' |
| | 'CAG repeats {gene}' |
| | 'heterozygous missense.{0,10}{gene}' |
| | 'heterozygous.*variant.{0,10}{gene}' |
| | 'sequence alteration .{0,10}{gene}' |
| | 'Gene panel show.{0,10}{gene} variant' |
| | 'Gene panel positive {gene}' |
| | '{gene}.[c]\.[0-9] .* VUS' |
| | 'Heterozygous VUS {gene}' |
| | 'variant {gene} gene identified' |
| | 'genetic mutation detected.*heterozygous.{0,10} {gene}' |
| | 'partial deletion {gene} gene, paternally inherited' |
| | 'Partial deletion {gene} gene, maternal inherited' |
| | 'Genetic test showed VUS {gene}' |

**Table S1.** Phrases used in the natural language processing algorithm to identify individuals with likely genetic epilepsy syndromes. Abbreviations: VUS – variant of unknown significance.

### Supplementary Table 2: Clinical characteristics of the study cohort

| ICD-10 | Likely genetic | Not genetic | Description |
|---|---|---|---|
| G40.419 | 51 | 9 | Other generalized epilepsy and epileptic syndromes, intractable, without status epilepticus |
| G40.319 | 37 | 20 | Generalized idiopathic epilepsy and epileptic syndromes, intractable, without status epilepticus |
| G40.824 | 31 | 12 | Epileptic spasms, intractable, without status epilepticus |
| G40.119 | 30 | 29 | Localization-related (focal) (partial) symptomatic epilepsy and epileptic syndromes without status epilepticus |
| G40.309 | 30 | 22 | Generalized idiopathic epilepsy and epileptic syndromes, not intractable, without status epilepticus |
| G40.802 | 24 | 22 | Other epilepsy, not intractable, without status epilepticus |
| G40.909 | 21 | 16 | Epilepsy, unspecified, not intractable, without status epilepticus |
| G40.911 | 21 | 3 | Epilepsy, unspecified, intractable, with status epilepticus |
| G40.109 | 20 | 17 | Localization-related (focal) (partial) symptomatic epilepsy and epileptic syndromes without status epilepticus |
| R56.9 | 19 | 16 | Unspecified convulsions |
| G40.209 | 18 | 10 | Localization-related (focal) (partial) symptomatic epilepsy and epileptic syndromes with complex partial seizures, not intractable, without status epilepticus |
| G40.901 | 18 | 10 | Epilepsy, unspecified, not intractable, with status epilepticus |
| R56.00 | 17 | 17 | Simple febrile convulsions |
| G40.822 | 15 | 6 | Epileptic spasms, not intractable, without status epilepticus |
| G40.814 | 14 | 1 | Lennox-Gastaut syndrome, intractable, without status epilepticus |
| G40.211 | 11 | 3 | Localization-related (focal) (partial) symptomatic epilepsy and epileptic syndromes with complex partial seizures, intractable, with status epilepticus |
| G40.409 | 11 | 3 | Other generalized epilepsy and epileptic syndromes, not intractable, without status epilepticus |
| F84 | 10 | 7 | Pervasive developmental disorders |
| G40.919 | 9 | 11 | Epilepsy, unspecified, intractable, without status epilepticus |
| R56.01 | 9 | 9 | Complex febrile convulsions |
| G40.301 | 8 | 3 | Generalized idiopathic epilepsy and epileptic syndromes, not intractable, with status epilepticus |
| G40.401 | 8 | 1 | Other generalized epilepsy and epileptic syndromes, not intractable, with status epilepticus |
| G40.42 | 8 | 0 | Cyclin-Dependent Kinase-Like 5 Deficiency Disorder |
| G40.834 | 8 | 0 | Dravet syndrome, intractable, without status epilepticus |
| G40 | 7 | 6 | Epilepsy and recurrent seizures |
| G40.A19 | 7 | 2 | Absence epileptic syndrome, intractable, without status epilepticus |
| G40.83 | 7 | 0 | Dravet syndrome |
| F84.0 | 6 | 6 | Autistic disorder |
| G40.201 | 6 | 6 | Localization-related (focal) (partial) symptomatic epilepsy and epileptic syndromes with complex partial seizures, not intractable, with status epilepticus |
| G40.219 | 6 | 6 | Localization-related (focal) (partial) symptomatic epilepsy and epileptic syndromes with complex partial seizures, intractable, without status epilepticus |
| R56.1 | 6 | 4 | Post traumatic seizures |
| G40.509 | 6 | 3 | Epileptic seizures related to external causes, not intractable, without status epilepticus |
| G40.801 | 6 | 3 | Other epilepsy, not intractable, with status epilepticus |
| G40.311 | 6 | 1 | Generalized idiopathic epilepsy and epileptic syndromes, intractable, with status epilepticus |
| G40.89 | 5 | 1 | Other seizures |
| G40.009 | 4 | 11 | Localization-related (focal) (partial) idiopathic epilepsy and epileptic syndromes with seizures of localized onset, not intractable, without status epilepticus |
| G40.A09 | 4 | 9 | Absence epileptic syndrome, not intractable, without status epilepticus |
| G40.001 | 4 | 3 | Localization-related (focal) (partial) idiopathic epilepsy and epileptic syndromes with seizures of localized onset, not intractable, with status epilepticus |
| F84.8 | 4 | 2 | Other pervasive developmental disorders |
| G40.011 | 4 | 2 | Localization-related (focal) (partial) idiopathic epilepsy and epileptic syndromes with seizures of localized onset, intractable, with status epilepticus |
| G40.821 | 4 | 2 | Epileptic spasms, not intractable, with status epilepticus |
| G40.823 | 4 | 1 | Epileptic spasms, intractable, with status epilepticus |

| | | | |
|---|---|---|---|
| F84.2 | 4 | 0 | Rett's syndrome |
| G40.803 | 3 | 4 | Other epilepsy, intractable, with status epilepticus |
| G40.411 | 3 | 0 | Other generalized epilepsy and epileptic syndromes, intractable, with status epilepticus |
| G40.501 | 2 | 2 | Epileptic seizures related to external causes, not intractable, with status epilepticus |
| G40.812 | 2 | 2 | Lennox-Gastaut syndrome, not intractable, without status epilepticus |
| G40.833 | 2 | 0 | Dravet syndrome, intractable, with status epilepticus |
| G40.B09 | 2 | 0 | Juvenile myoclonic epilepsy, not intractable, without status epilepticus |
| G40.111 | 1 | 2 | Localization-related (focal) (partial) symptomatic epilepsy and epileptic syndromes with simple partial seizures, intractable, with status epilepticus |
| G40.019 | 1 | 1 | Localization-related (focal) (partial) idiopathic epilepsy and epileptic syndromes with seizures of localized onset, intractable, without status epilepticus |
| G40.B19 | 1 | 1 | Juvenile myoclonic epilepsy, intractable, without status epilepticus |
| G40.10 | 1 | 0 | Localization-related (focal) (partial) symptomatic epilepsy and epileptic syndromes with simple partial seizures, not intractable |
| G40.31 | 1 | 0 | Generalized idiopathic epilepsy and epileptic syndromes, intractable |
| G40.811 | 1 | 0 | Lennox-Gastaut syndrome, not intractable, with status epilepticus |
| G40.A01 | 1 | 0 | Absence epileptic syndrome, not intractable, with status epilepticus |
| F84.5 | 0 | 1 | Asperger's syndrome |

**Table S2.** The epilepsy syndromes associated with the participants from the matched case-control cohort (n = 503) are shown as ICD-10 codes for the case ("Likely genetic") and control ("Not genetic") groups, respectively, sorted by overall prevalence. Codes are shown if they are grouped under G40 ("Epilepsy and recurrent seizures"), R56 ("Convulsions, not elsewhere classified"), or F84 ("Pervasive developmental disorders). Note that individuals may be assigned more than one ICD-10 code over the duration in the healthcare system.
